# The Separate and Joint Associations of Own and Spousal Depression with Mortality in Couples

**DOI:** 10.1101/2025.09.15.25335774

**Authors:** Toshiaki Komura, Säde Stenlund, Laura D Kubzansky

**Author notes:** **Corresponding author:** Toshiaki Komura Department of Social and Behavioral Sciences, Harvard T. H. Chan School of Public Health, Boston, MA, USA.

## Abstract

**Background:** Depression causes a substantial burden on a person’s health and well-being. However, evidence is limited regarding whether depression of one person in a marital relationship may affect the other partner’s health. This study examined whether depression in one partner within a married couple is associated with the other partner’s mortality, also considering whether only one or both partners have depression.

**Methods:** To test the hypothesis that spousal depression is associated with higher mortality risk for their partner, we used a nationally representative sample of older US adults from the Health and Retirement Study. Depression was assessed in both partners using the Center for Epidemiologic Studies Depression Scale. With baseline measures in 2010, we characterized depression status for 8,442 married individuals within 4,225 couples as follows: i) no depression in either partner, ii) depression in respondent only, iii) depression in spouse only, and iv) depression in respondent and their spouse. Associations between couples’ depression status and individuals’ mortality over 11 years were estimated using Cox proportional hazards models adjusted for 22 covariates.

**Results:** Higher mortality hazard was observed when only the respondent had depression (hazard ratio (HR): 1.44 [95% confidence interval (CI): 1.26, 1.63]), but also when only their spouse exhibited depression (HR: 1.18 [95% CI: 1.00, 1.38]). When both partners had depression, a jointly elevated mortality hazard was observed (HR: 1.56 [95% CI: 1.30, 1.87]).

**Conclusions:** Results suggest the harmful effects of depression could extend beyond the individual to one’s partners’ physical health. Future studies on health effects of depression should incorporate familial contexts.

**Highlights:**

- When one partner was depressed, the other partner had a higher mortality risk regardless of their own depression status.
- Widowhood effect did not fully explain the effects of spousal depression on one’s mortality.
- Harmful effects of depression could extend beyond the individual to the partners’ physical health.

## Introduction

Depression has been linked to multiple adverse health consequences beyond mental health and is linked to higher mortality risks with several underlying mechanisms posited (Zhang et al., 2023). For example, depression could lead to unhealthy lifestyles, such as sleep disturbance and less physical activity (Ford et al., 2012; M. J. Murphy & Peterson, 2015; Roshanaei-Moghaddam et al., 2009). Alongside these behavioral factors, depression may also trigger recurring biological stress responses, including dysfunction in the autonomic nervous system and immune inflammation, that in turn elevate risks for chronic disease and premature mortality (Hare et al., 2014; Nemeroff & Goldschmidt-Clermont, 2012; Udupa et al., 2007). Moreover, having depression can increase other psychosocial problems, such as social isolation, which have in turn been identified as risk factors for mortality (Luo, 2023). Given the substantial and robust findings of depression with risk of adverse health and mortality, an important question is whether effects of a person’s depression might extend beyond their own health to other individuals in intimate relationships, particularly with regard to potential effects on physical health. Although some work has suggested that psychological factors in one individual can affect the physical health of close partners (e.g., one partner’s optimism is associated with higher cognitive function of the other partner) (Oh et al., 2020), no study has examined whether and the extent to which a person’s depression may affect their partner’s mortality. Evaluating the effect of one person’s depression on their partner’s health comes with methodological challenges, particularly with regard to assessing causality and direction of effects. These include inability not only to randomize individuals to be depressed or not for a long period of time but also to randomize marital partners. A key concern is assortative mating, where individuals are more likely to be married to those who share similar traits (Robinson et al., 2017). Such traits could drive any apparent similarity in health and well-being over the life course, making it difficult to assess unique effects of other factors that come into play during the marriage.

Animal research examining effects of induced stress within mated animals, suggests transmission of health risk across partners is possible. An experimental study with zebra finches reported an intriguing effect of stress on mortality risk within mated birds (Monaghan et al., 2012). The birds were randomized to experience early life stress or not and then subsequently randomly assigned to mates who either did or did not also receive the early life stress exposure (Monaghan et al., 2012). These experimental treatments resulted in four exposure groups: i) only the male bird was stress exposed, ii) only the female bird was stress exposed, iii) both birds in the dyad were stress exposed, and iv) neither bird in the dyad was stress exposed (Monaghan et al., 2012). Over a 3 year follow-up period, stress exposed birds who mated with birds who were also stress exposed showed the highest, and stress unexposed birds mated with stress unexposed birds had the lowest mortality rates (Monaghan et al., 2012). Of particular interest, stress unexposed birds mated with stress exposed birds had a higher risk of mortality compared to that of birds who were both stress unexposed and as high as birds who were themselves stress exposed but whose mates were not (Monaghan et al., 2012). These provocative findings suggest that one partner’s stress or distress could be transmitted and affect the other partner’s health. Such effects might be particularly evident in a close relationship like a marriage and in older adulthood when individuals are increasingly vulnerable to poor health.

This study aimed to investigate the separate and joint associations of depression on mortality in partners within a couple. We tested the hypothesis that individuals whose partners are depressed will exhibit increased mortality risk. We used data from a nationally representative survey of US adults aged ≥50 from the Health and Retirement Study (HRS). We included a broad range of covariates encompassing sociodemographic characteristics and physical health conditions of each partner to address concerns about potential confounding associated with assortative mating to the greatest extent possible (Robinson et al., 2017). Moreover, given prior work showing depression elevates mortality risks, and that experiencing the death of a spouse also increases risk of depression and mortality (also called the widowhood effect) (Elwert & Christakis, 2008; Moon et al., 2011; Zhang et al., 2023). partner bereavement could be one pathway by which having a depressed spouse could lead to an individual exhibiting increased risk of mortality. Hence, we also considered whether the association between one spouse’s mortality risk and the other spouse’s depression might vary before and after widowhood.

## Methods

### Population

Data are from HRS, a nationally representative sample of US adults aged ≥50. HRS is an ongoing panel study that started in 1992 and conducts biannual surveys via in-person interviews, telephone, or mailed questionnaires. HRS recruited 12,652 participants (index individuals) in the first wave and has added new samples in every following wave. For married participants, HRS also collects information from their spouses. Further details of the cohort profile can be found elsewhere (Sonnega et al., 2014).

We used data from the earliest possible waves based on when depression and key covariates were all assessed, resulting in 2008 serving as the pre-baseline to measure covariates that may confound the associations of interest and 2010 serving as baseline, when exposure status was assessed (i.e., couple’s depression status). Use of pre-baseline data ensures appropriate temporality between exposure and covariates to minimize the unintended adjustment of potential mediators in analyses (VanderWeele et al., 2020). Of the 18,665 index individuals (i.e., participants) from HRS 2008, we excluded index individuals who met any of the following criteria: i) self-reported unmarried status (including divorced, widowed, or separated) in HRS 2008, ii) spouse did not participate the HRS 2008 survey, iii) spouse did not report as married even though the index individual did, iv) index individuals or their spouses did not participate in the HRS 2010 survey (i.e., exposure status was unidentifiable), and v) identical spouses from the pre-baseline did not participate in the HRS 2010 survey (i.e., covariate information of the spouse was unmeasured). We also excluded 8 index individuals because their recorded death dates were prior to the interview date at baseline, and therefore, likely to be misrecorded. After applying these eligibility criteria, we included 8,442 individuals from 4,225 couples (see **Figure S1** for sample inclusion flow chart).

All index individuals were followed from 2010 until any of the following events occurred: i) death of the index individual (i.e., outcome), ii) censoring (i.e., loss to follow-up), and iii) administrative censoring in May 2021 (end of the study period). For couples with both partners included in the analyses, an individual was treated as i) an index individual, and ii) the spouse of another index individual from the same household.

### Exposure

Depression in index individuals and their spouses was assessed at the study baseline (HRS 2010) using the validated 8-item Center for Epidemiologic Studies Depression (CES-D8) scale (Radloff, 1977). Each participant and their spouse were asked dichotomous (yes=1/no=0) questions about whether they felt any of the 8 experiences much of the time during the past week (e.g., “felt depressed,” and “sleep was restless”). A total score ranging from 0 to 8 was calculated by computing the sum of these items, where higher scores indicate more depressive symptoms.

We defined index individuals or their spouses as having probable depression using a previously validated cutpoints of CES-D8 score ≥3 (Steffick, 2000). Subsequently, all individuals were classified into 4 exposure groups: 1) no depression in the either partner, 2) depression in the index individual only, 3) depression in the spouse only, and 4) depression in both. Index individuals in exposure group 1 (no depression in either partner) were treated as the reference in all analyses. Missing items in CES-D8 items were imputed via multiple imputation.

### Outcome

The outcome was all-cause mortality over 11 years of follow-up. HRS collects information of death from an exit interview conducted with next-of-kin and a search of the National Death Index database following the interview. The mortality of index individuals was assessed starting after the baseline interview in 2010 through May 2021, the latest date on which HRS last collected mortality information. The date of death is recorded on a monthly basis.

### Covariates

We selected 22 characteristics of index individuals, their spouses, and the household as potential confounders based on prior work (Gilman et al., 2017; R. A. Murphy et al., 2016). These covariates included sociodemographic characteristics (age, gender, race, education, religious denomination, employment, health insurance, and a childhood adversity score), physical health conditions and disability (a comorbidity score summarizing history four major chronic conditions, overweight/obesity, cognitive impairment/dementia, and an instrumental activities of daily living (IADL) score), a composite health behavior score, and household characteristics (income, wealth, and length of marriage). **Appendix** describes the covariates included in the analysis. **Table S1** describes further details about how each covariate was coded from the original questionnaires.

### Statistical analysis

We first assessed the distribution of covariates across exposure subgroups. Second, we examined associations between each couple’s profile of depression and the index individuals’ mortality by plotting Kaplan-Meier curves of the 4 exposure subgroups. Third, Cox proportional hazards models were applied to estimate hazard ratios (HRs) of index individuals’ mortality associated with the profile of depression status in couples. Because our analytic sample included index individuals from the same couple, models included random intercept for each household to account for potential clustered correlation. To assess the extent to which potential confounding from specific types of covariates might affect associations of interest, we conducted four increasingly adjusted models. We first ran an unadjusted model (Model 1). Model 2 included sociodemographic covariates (age, gender, race, education, religious denomination, employment, health insurance, childhood adversity score, income, wealth, and length of current marriage). Model 3 further added physical health conditions and disability (comorbidity score, overweight/obesity, cognitive impairment/dementia, and IADL score). Model 4 further adjusted respondent’s health behavior score. A proportional hazard test of Model 4 by Schoenfeld residuals showed a p-value of 1.00, indicating the proportional hazard assumption is unlikely to be violated.

We performed the following sensitivity analyses. First, since 1,650 participants were excluded from the analytic sample at the baseline due to the dropout or participation of a new spouse, the selection of the sample occurring between pre-baseline and baseline might cause bias. We thus repeated the survival analysis further adjusting for all covariates and the inverse probability of censoring weighting (IPCW) to understand the extent to which this potential bias might change our findings (Dong et al., 2020). Second, because multiple imputation predicting depressive symptoms of those who did not report the majority of the items requires stronger assumptions, we conducted analyses instead using mean imputation for CES-D8 scores if participants had responded to half or more of the items (Demissie et al., 2003). In this analysis, 7,296 (73.4%) index individuals were included. Third, because HRS collects data via proxy responses from household members when the primary participants cannot respond to the survey, we ran an analysis without those who had proxy responses at the pre-baseline. In our analytic sample, covariate information for 426 participants (5.0%) was measured by proxy interviews, but CES-D8 item was not. This analysis included 8,016 index individuals.

Finally, we performed analyses using multistate models to investigate whether spousal death plays a role in any observed relationship between depression status profiles and index respondents’ mortality hazard. Briefly, multistate modeling is an extension of Cox proportional hazards model techniques that can estimate HRs across multiple states that could occur during follow-up, including competing risks and intermediate events (Aalabaf-Sabaghi, 2019). In our setting, we first defined a spouse’s death during the follow-up as an intermediate event. We then estimated HRs associated with each couple’s depression status profile during the following periods: i) before a spouse’s death and ii) after a spouse’s death. The analysis is based on an additional assumption of Markov property, which suggests that mortality hazard in a given month during the follow-up is determined solely by whether spousal death already occurred at that time but not take account of the duration of time since the spouse’s death (if occurred). Because we additionally incorporated information about the spouse’s death in this multistate analysis, 10 index individuals whose spouses were deceased or lost to follow-up before the baseline interview were excluded, resulting in 8,432 index individuals. Index individuals were followed until any of the following events occurred: i) death of the index individual, ii) censoring, iii) administrative censoring in May 2021, iv) censoring of their spouses not due to death (i.e., the intermediate status became unidentifiable). **Figure S2** illustrates a conceptual diagram and the distributions of index individuals that experienced each state.

Because HRS uses a complex multistage probability survey design, we applied sampling weights in all analyses. Except when otherwise indicated, all missing covariates and depressive symptom items were imputed via multiple imputation using R package *mice* (*Multivariate Imputation by Chained Equations [R package mice version 3.16.0]*, 2023). Missingness ranged from 0% to 15.1% with the largest proportion of missing data in childhood adversity score. Imputation for at least one CES-D8 item was performed among 13.6% index individuals. All analyses were performed using 10 imputed samples, and their estimates were combined based on Rubin’s rule (Rubin, 2004). All analyses were conducted using R version 4.3.1. The present study used publicly available data that collected informed consent from all participants. Thus, the Harvard T.H. Chan School of Public Health IRB exempted it from review.

## Results

### Baseline characteristics of the study population

**Table 1** shows the distribution of pre-baseline covariates within 8,442 index individuals (4,225 couples) and across four depression status profiles. Depression status profiles included 3.5% with both partners depressed, 9.1% each with only index individual or spouse depressed, and the largest percentage (64.8%) for neither partner having probable depression. The mean age (standard deviation [SD]) was 66.52 (9.34). Compared to the reference group, index individuals with depression themselves or whose spouses were depressed were more likely to be of non-White race/ethnicity. Furthermore, these individuals were more likely to have disadvantaged socioeconomic status, to have a history of chronic diseases, and to engage in unhealthy behaviors. The most disadvantaged sociodemographic conditions and poorer health were observed in index individuals who were depressed and who also had depressed partners, compared to all other depression status groups.

**Table 1:** Characteristics of 8,442 index individuals according to their depression status profile.^*,†^.

| Characteristics, n (%) | All Sample | Depression in |  |  |  |
| --- | --- | --- | --- | --- | --- |
|  |  | No Depression | Index Individual Only | Depression in Spouse Only | Depression in Both |
| Individuals, No Death | N = 8,442<br>2,498 (30%) | N = 5,468<br>1,377 (25%) | N = 765<br>279 (36%) | N = 765<br>244 (32%) | N = 298<br>123 (41%) |
| **Sociodemographic Factors** |  |  |  |  |  |
| Age, mean (SD) | 66.52 (9.34) | 66.42 (8.99) | 65.47 (9.76) | 66.24 (9.93) | 64.70 (10.03) |
| Age of Spouse, mean (SD) | 66.52 (9.34) | 66.42 (8.99) | 66.25 (9.93) | 65.47 (9.75) | 64.72 (10.10) |
| Gender |  |  |  |  |  |
| Male | 4,220 (50%) | 2,734 (50%) | 282 (37%) | 483 (63%) | 149 (50%) |
| Female | 4,222 (50%) | 2,734 (50%) | 483 (63%) | 282 (37%) | 149 (50%) |
| Race |  |  |  |  |  |
| White | 6,683 (79%) | 4,497 (82%) | 561 (73%) | 563 (74%) | 210 (70%) |
| Black | 751 (8.9%) | 457 (8.4%) | 82 (11%) | 81 (11%) | 31 (10%) |
| Hispanic/Latino | 809 (9.6%) | 406 (7.4%) | 97 (13%) | 94 (12%) | 50 (17%) |
| Other Identity | 198 (2.3%) | 108 (2.0%) | 25 (3.3%) | 26 (3.4%) | 7 (2.3%) |
| Missing | 1 | 0 | 0 | 1 | 0 |
| Race of Spouse |  |  |  |  |  |
| White | 6,684 (79%) | 4,498 (82%) | 563 (74%) | 561 (73%) | 210 (70%) |
| Black | 751 (8.9%) | 457 (8.4%) | 81 (11%) | 82 (11%) | 31 (10%) |
| Hispanic/Latino | 808 (9.6%) | 405 (7.4%) | 94 (12%) | 97 (13%) | 50 (17%) |
| Other Identity | 198 (2.3%) | 108 (2.0%) | 26 (3.4%) | 25 (3.3%) | 7 (2.3%) |
| Missing | 1 | 0 | 1 | 0 | 0 |
| Education |  |  |  |  |  |
| <High School | 1,730 (21%) | 879 (16%) | 222 (29%) | 196 (26%) | 114 (38%) |
| High School | 2,581 (31%) | 1,699 (31%) | 224 (29%) | 221 (29%) | 79 (27%) |
| >High School | 4,128 (49%) | 2,888 (53%) | 318 (42%) | 348 (45%) | 105 (35%) |
| Missing | 3 | 2 | 1 | 0 | 0 |
| Education of Spouse |  |  |  |  |  |
| <High School | 1,731 (21%) | 880 (16%) | 196 (26%) | 222 (29%) | 114 (38%) |
| High School | 2,580 (31%) | 1,699 (31%) | 221 (29%) | 224 (29%) | 79 (27%) |
| >High School | 4,128 (49%) | 2,887 (53%) | 348 (45%) | 318 (42%) | 105 (35%) |
| Missing | 3 | 2 | 0 | 1 | 0 |
| Religious denomination |  |  |  |  |  |
| Protestant | 5,311 (63%) | 3,536 (65%) | 456 (60%) | 456 (60%) | 175 (59%) |
| Catholic | 2,307 (27%) | 1,426 (26%) | 231 (30%) | 219 (29%) | 95 (32%) |
| Jewish | 153 (1.8%) | 86 (1.6%) | 16 (2.1%) | 12 (1.6%) | 11 (3.7%) |
| Other Identity | 86 (1.0%) | 54 (1.0%) | 7 (0.9%) | 15 (2.0%) | 0 (0%) |
| None | 557 (6.6%) | 351 (6.4%) | 52 (6.8%) | 61 (8.0%) | 15 (5.1%) |
| Missing | 28 | 15 | 3 | 2 | 2 |
| Religious |  |  |  |  |  |
| denomination of Spouse |  |  |  |  |  |
| Protestant | 5,311 (63%) | 3,536 (65%) | 456 (60%) | 456 (60%) | 175 (59%) |
| Catholic | 2,307 (27%) | 1,426 (26%) | 219 (29%) | 231 (30%) | 95 (32%) |
| Jewish | 153 (1.8%) | 86 (1.6%) | 12 (1.6%) | 16 (2.1%) | 11 (3.7%) |
| Other Identity | 86 (1.0%) | 54 (1.0%) | 15 (2.0%) | 7 (0.9%) | 0 (0%) |
| None | 557 (6.6%) | 351 (6.4%) | 61 (8.0%) | 52 (6.8%) | 15 (5.1%) |
| Missing | 28 | 15 | 2 | 3 | 2 |
| Household Income (thousand dollars), mean (SD) | 84.69 (113.48) | 92.64 (123.94) | 68.06 (71.14) | 68.06 (71.14) | 50.85 (61.22) |
| Household Wealth (thousand dollars), mean (SD) | 655.12 (1,289.10) | 717.56 (1,371.35) | 517.10 (989.72) | 517.10 (989.72) | 292.43 (590.31) |
| Employment |  |  |  |  |  |
| Not Employed | 871 (10%) | 457 (8.4%) | 142 (19%) | 64 (8.4%) | 70 (23%) |
| Retired | 4,016 (48%) | 2,574 (47%) | 358 (47%) | 355 (46%) | 133 (45%) |
| Employed | 3,555 (42%) | 2,437 (45%) | 265 (35%) | 346 (45%) | 95 (32%) |
| Employment of Spouse |  |  |  |  |  |
| Not Employed | 868 (10%) | 457 (8.4%) | 64 (8.4%) | 142 (19%) | 70 (23%) |
| Retired | 4,020 (48%) | 2,575 (47%) | 355 (46%) | 358 (47%) | 133 (45%) |
| Employed | 3,554 (42%) | 2,436 (45%) | 346 (45%) | 265 (35%) | 95 (32%) |
| Health Insurance (yes) | 8,022 (95%) | 5,267 (96%) | 711 (93%) | 708 (93%) | 256 (86%) |
| Missing | 5 | 2 | 0 | 0 | 0 |
| Health Insurance of Spouse (yes) | 8,022 (95%) | 5,267 (96%) | 711 (93%) | 708 (93%) | 256 (86%) |
| Missing | 3 | 1 | 0 | 0 | 0 |
| Length of Current Marriage, Years, (SD) | 36.76 (15.80) | 36.71 (15.54) | 35.35 (16.45) | 35.43 (16.32) | 33.46 (16.37) |
| Missing | 176 | 90 | 19 | 21 | 13 |
| Childhood Adversity Score, mean (SD)<sup>‡</sup> | 0.93 (0.96) | 0.92 (0.95) | 0.98 (1.02) | 0.92 (0.98) | 0.96 (0.99) |
| Missing | 1273 | 573 | 124 | 126 | 73 |
| **Physical Health** |  |  |  |  |  |
| Comorbidity Score<sup>§</sup> |  |  |  |  |  |
| Index Individual | 0.61 (0.79) | 0.56 (0.75) | 0.72 (0.84) | 0.65 (0.80) | 0.78 (0.89) |
| Spouse | 0.61 (0.79) | 0.56 (0.75) | 0.65 (0.80) | 0.72 (0.84) | 0.78 (0.88) |
| Overweight/obesity |  |  |  |  |  |
| Index Individual | 6,036 (72%) | 3,880 (72%) | 570 (76%) | 593 (78%) | 215 (73%) |
| Missing | 106 | 49 | 17 | 6 | 5 |
| Spouse | 6,035 (72%) | 3,880 (72%) | 593 (78%) | 570 (76%) | 215 (73%) |
| Missing | 105 | 49 | 6 | 17 | 5 |
| Cognitive Impairment/Dementia<sup>¶</sup> |  |  |  |  |  |
| Index Individual | 1,196 (14%) | 606 (11%) | 153 (20%) | 134 (18%) | 91 (31%) |
| Missing | 426 | 65 | 16 | 21 | 7 |
| Spouse | 1,198 (14%) | 606 (11%) | 134 (18%) | 153 (20%) | 93 (31%) |
| Missing | 426 | 65 | 16 | 21 | 7 |
| **IADL Score\*\*** |  |  |  |  |  |
| Index Individual | 0.17 (0.63) | 0.08 (0.39) | 0.36 (0.83) | 0.13 (0.50) | 0.45 (0.92) |
| Missing | 1 | 1 | 0 | 0 | 0 |
| Spouse | 0.17 (0.63) | 0.08 (0.39) | 0.13 (0.50) | 0.36 (0.83) | 0.46 (0.93) |
| Missing | 1 | 1 | 0 | 0 | 0 |
| **Health Behaviors** |  |  |  |  |  |
| **Health Behavior Score<sup>††</sup>** |  |  |  |  |  |
| Index Individual | 0.61 (0.68) | 0.56 (0.66) | 0.73 (0.70) | 0.67 (0.70) | 0.84 (0.74) |
| Missing | 124 | 67 | 11 | 10 | 5 |
| Spouse | 0.61 (0.68) | 0.56 (0.66) | 0.73 (0.70) | 0.67 (0.70) | 0.84 (0.74) |
| Missing | 123 | 67 | 11 | 10 | 5 |

Abbreviations: IADL score, instrumental activities of daily living score.
\*Data are reported n (%) for categorical variables, mean (SD) for continuous variables.
†Exposure subgroups include only those with a complete CES-D8 score at the baseline because those with missing CES-D8 item could be classified in different exposure subgroups after multiple imputation. Those whose CES-D8 items were imputed are included only in the column “All Sample.”
‡Childhood adversity score was measured using the composite sum score of four items (yes/no; 1 point assigned for presence of each adversity): 1) physical abuse, 2) alcohol/drug issue of parents, 3) repeating a year of school, and 4) trouble with police.
§Comorbidity score was measured based on the composite sum score of four disease histories (yes/no; 1 point assigned for presence of each disease history): 1) heart disease, 2) stroke, 3) diabetes, and 4) cancer.
¶Cognitive impairment/dementia was assessed using the Telephone Interview for Cognitive Status (TICS, 0-27 points). Those with TICS score <12 were classified as having cognitive impairment/dementia.
\*\*IADL score was derived from the composite sum score of 5 items (yes/no; 1 point assigned for presence of difficulty with each activity): 1) use telephone, 2) managing money, 3) take medications, 4) shop for grocery, and 5) prepare hot meal.
††Health behavior score was measured based on the composite sum score of three unhealthy behaviors (yes/no; 1 point assigned for presence of each unhealthy behavior): 1) binge drinking, 2) smoking, and 3) physical inactivity.

### Depression status in couples and index individuals’ mortality

Over the median follow-up period of 113 months, 2,498 index individuals (29.6%) died. Individuals in couples where either the index individual or their spouse had depression also showed higher mortality over the follow-up period compared to individuals in couples where neither partner had depression at baseline (see Kaplan-Meier curve; **Figure 1**).

**Figure 1.**
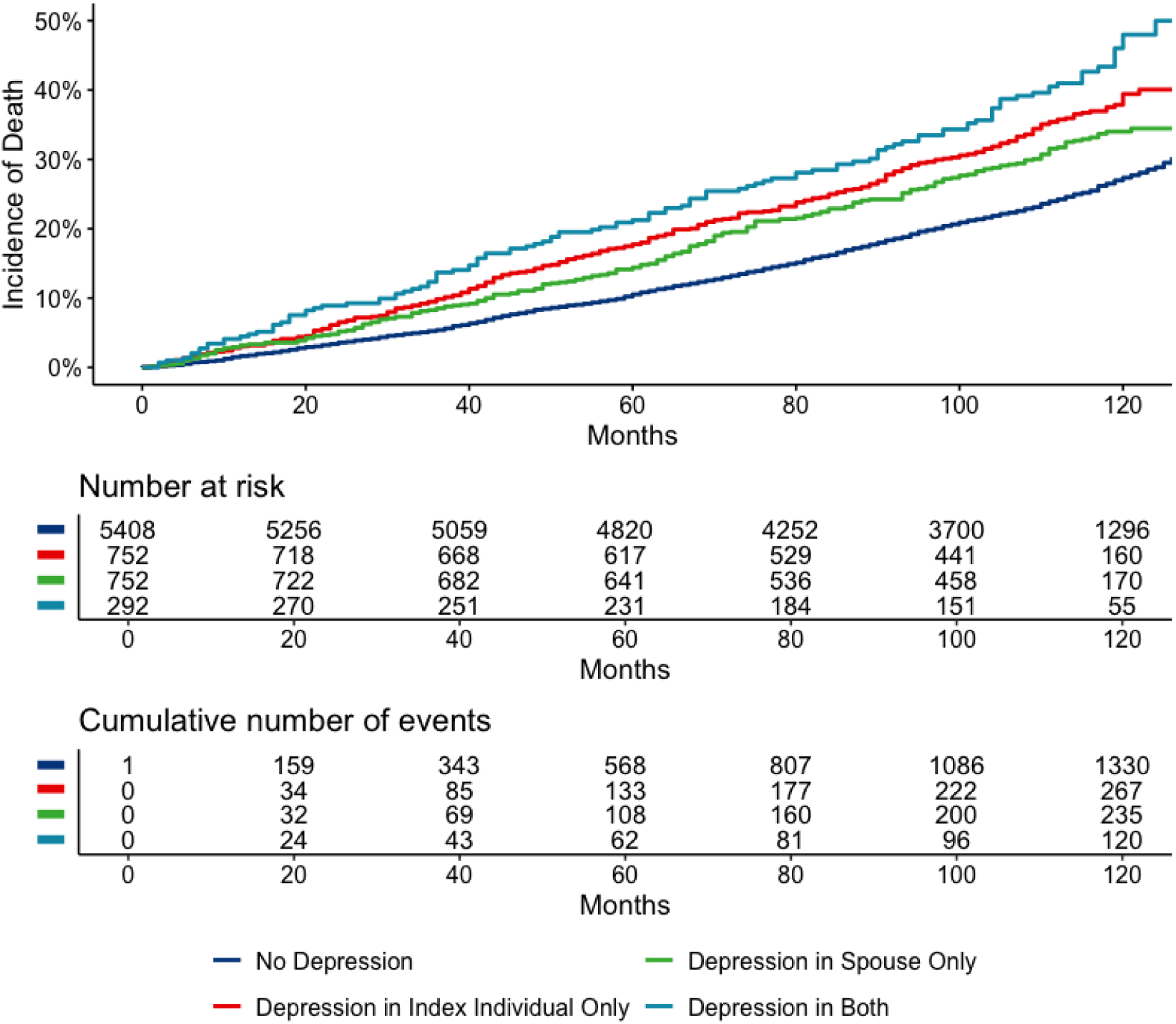
Kaplan-Meier Curve of Index Individuals’ Death by Depression Status Profile (N = 7,296).* *The tables show the number at risk and cumulative number of events among the exposure subgroups, only including those with a complete CES-D8 score at the baseline, because those with missing CES-D8 item could be classified in different exposure subgroups after multiple imputation.

During the follow-up period, compared to individuals in couples where neither partner reported depression, unadjusted mortality hazard was higher for index individuals in couples with only one depressed partner (**Table 2**). With the same reference group, index individuals in couples where both spouses had depression had the highest mortality hazard. Overall trends remained evident after adjusting for all covariates (depression_index only_: HR [95% confidence interval (CI)] = 1.44 [1.26, 1.63]; depression_spouse only_: HR [95% CI] = 1.18 [1.00, 1.38]; depression_both_: HR [95% CI] = 1.56 [1.30, 1.87]).

**Table 2.**
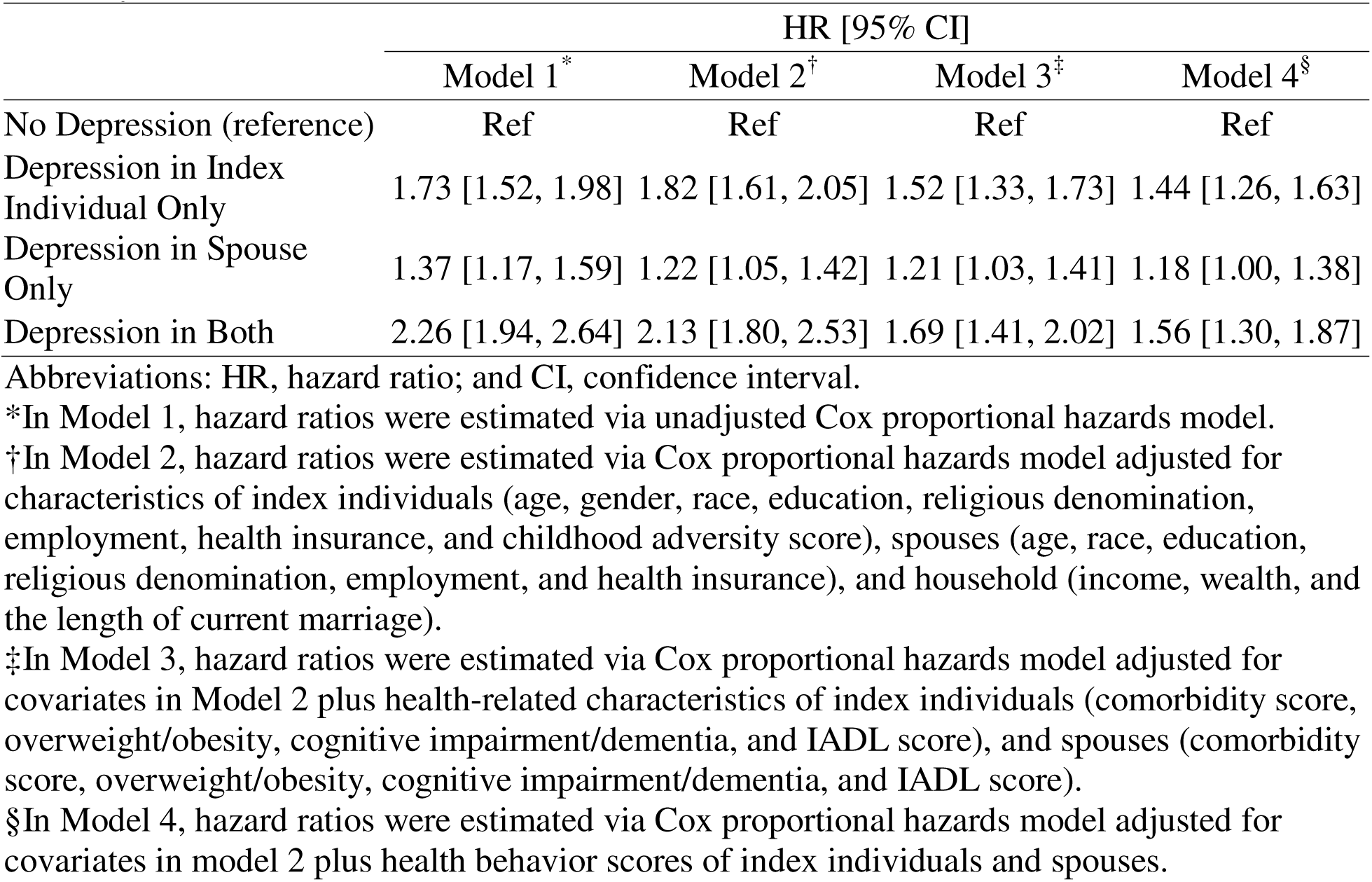
Associations between Depression Status Profile in Couples and Index Individual’s Mortality (All Models, N = 8,442).

Findings from sensitivity analyses using censoring weights to account for individuals who were eligible for inclusion in the study at pre-baseline but excluded at baseline due to drop out showed widened confidence intervals that included null values for all exposure subgroups (**Figure S3**). In the sensitivity analysis with CES-D8 items imputed only among respondents who completed at least the half of the questionnaires rather than imputing all items regardless of how many were completed, mortality hazards associated with depression status in couples were comparable to the main analysis, although the confidence interval for couples where only spouse was depressed was widened to include the null value (**Figure S4**). An analysis excluding individuals with covariates measured by proxy responses yielded a similar trend as the main analyses (**Figure S5**).

### Mortality associated with depression in couples before and after widowhood

Multistate model analyses demonstrate elevated HRs for death associated with a couple’s depression status profile before compared to after a spouse’s death (**Figure 3**). Before their spouses was deceased, index individuals in couples with one versus no depressed partner(s) had higher HRs of mortality (depression_index only_: HR [95% CI] = 1.36 [1.11, 1.66]; depression_spouse only_: HR [95% CI] = 1.30 [1.07, 1.57]; with highest hazard evident among index individuals in couples with both partners experiencing depression: HR [95% CI] = 1.54 [1.18, 2.02]). On the other hand, HRs for death after versus before widowhood were generally smaller in all exposure subgroups.

**Figure 3.**
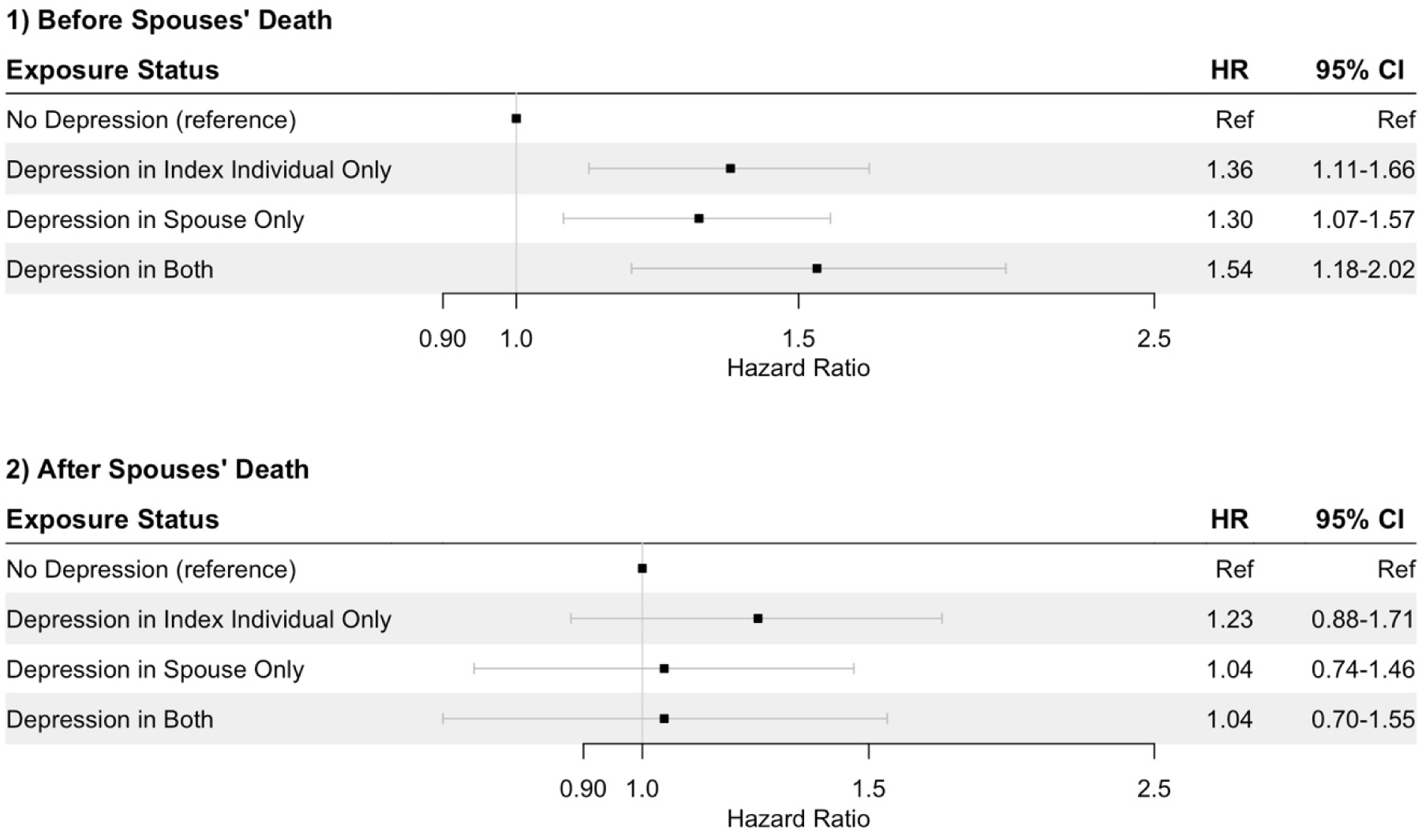
Multistate Model Analysis on Potential Pathways between Depression in Couples and Mortality (N = 8,432).* Abbreviations: HR, hazard ratio; and CI, confidence interval. *Hazard ratios were estimated via Multistate model adjusted for characteristics of index individuals (age, race, education, religious denomination, employment, health insurance, and childhood adversity score, comorbidity score, overweight/obesity, cognitive impairment/dementia, instrumental activities of daily living score, and health behavior score), spouses (age, race, education, religious denomination, employment, health insurance, comorbidity score, overweight/obesity, cognitive impairment/dementia, instrumental activities of daily living score, and health behavior score), and household (income, wealth, and the length of current marriage).

## Discussion

It has long been appreciated that mental health can affect physical health and that mental health of one partner can affect that of the other. However, findings from this study suggest that effects of mental health may have an even longer reach, beyond the individual’s own physical health to that of their close partners. This prospective longitudinal study of older married adults demonstrates that depression in one spouse is associated with a greater mortality hazard of their partner regardless of the index individual’s own depression level. First, we observed that index individuals’ own and spousal depression were separately associated with elevated mortality hazard for the individual. Second, we observed jointly elevated mortality hazards when both individuals in the couple had depression. Our multistate model analysis also found spousal depression was associated with higher hazards of mortality in index individuals before widowhood only.

Most sensitivity analyses showed consistent trends. The models with IPCW to accout for the dropout at the baseline showed the interval estimates of mortality hazards that included null values in all exposure subgroups. However, we believe this result is likely due to the lack of statistical power, and therefore should not completely negate our findings, given that relationships between one’s own depression and mortality have been well-established, and weighting approaches often come with higher variance (Ellis & Brookhart, 2013; Zhang et al., 2023). Furthermore, the mortality hazard associated with spousal depression only included the null value when the mean imputation was applied, while the point estimate was consistent across the main analyses. The results might suggest instability in our main findings due to exposure classification. At the same time, the result might be affected by the lack of statistical power due to small sample sizes in exposure subgroups.

Most work linking depression with poor physical health has focused on an individual’s experience of depression and how that affects their later health (Hawton et al., 2013; Kraus et al., 2019; Tunvirachaisakul et al., 2018). The impact that spousal depression might have on the health of their partner has received limited attention. The experiment examining effects of early life stress on longevity using zebra finche dyads discovered that high stress levels in one mate negatively affected the other mate’s risk of mortality with the effect magnitude for having a stressed mate appearing similar to that of experiencing stress directly (Monaghan et al., 2012). Remarkably, our study showed strikingly similar patterns among human marital partners considering profiles of depression in the couple (Monaghan et al., 2012). Of note, while the birds were randomly assigned to mate with partners that were or were not stressed, in the current study, we assume most individuals chose their partners intentionally. Thus, a major concern in our setting is the potential bias induced by assortative mating, whereby couples might share similar traits including with regard to health and longevity (Robinson et al., 2017). Importantly, assortative mating itself does not directly induce individuals’ death; rather it suggests that unhealthy people are more likely to marry other unhealthy people with both partners exhibiting poor health later on due to initial health status, regardless of spousal mental health. We thus adjusted our analyses with a broad range of major characteristics of index individuals, spouses, and households that may be associated with both depression and death to mitigate this concern. Notably, even after adjusting for all covariates, our findings showed substantial associations with increased mortality hazard (ranging from 10 to 34 % higher) among non-depressed individuals whose partners did versus did not have depression. The magnitude of mortality hazard associated with spousal depression in the current study is similar to findings of elevated mortality risk with other known risk factors such as sedentary behavior or an unhealthy diet (Chau et al., 2015; Gao et al., 2021).

Prior work on excess risk of mortality associated with depression has identified potential mechanisms underlying these associations (Pan et al., 2011; Penninx et al., 1999; Pratt et al., 2016; Zhang et al., 2023), and some of these mechanisms may be at play in associations linking excess mortality with having a depressed spouse. Prior work has demonstrated depressed individuals have higher likelihood of engaging in unhealthy behaviors, including physical inactivity and increased alcohol consumption (Bonnet et al., 2005; Cabello et al., 2017). One’s health behaviors could also be influenced by their partner’s behaviors (Cornelius et al., 2016; Kiecolt-Glaser & Wilson, 2017). Similarly, depression could also cause an imbalance in interdependence in daily functioning and other activities, such as cooking (resulting in deteriorated quality of nutrition intake), caring for other family members thereby imposing increased burden and stress on the partner, and perhaps greater biological stress responses. Moreover, individuals in couples where both spouses had depression showed the highest mortality hazards. Social support from relationships, including family and friends, is a crucial determinant of recovery and subjective well-being of individuals with depression (Kuehner & Buerger, 2005; Wickramaratne et al., 2022). However, such buffering function of the family might be hindered when both persons in a couple suffer from depression. Further research is needed to explore whether and the degree to which these potential mechanisms account for the separate and joint associations between depression in a couple and an individual’s mortality hazard.

Intriguingly, our multistate model analysis showed associations between spousal depression and higher hazards of mortality in index individuals even before the spouse died. This finding suggests an individual’s mortality could be affected by spouses’ depression whether or not their spouses died over the course of follow-up. However, our findings suggest the widowhood effect does not fully explain the effects of spousal depression on index individuals’ mortality. In fact, associations between spousal depression and index individuals’ hazard of mortality were substantially weaker after widowhood, which may suggest the impact of widowhood on mortality overrides any hazard associated with their spouse’s depression prior to death.

Our findings have important implications for public health, clinical practice, and research. First, while most applied research discusses the health effects of depression at the individual level, our results suggest health effects of depression may go beyond the individual directly experiencing depression (Kuehner & Buerger, 2005; Wickramaratne et al., 2022). Therefore, future research on the health impact of depression needs to account for intimate relationships and psychosocial functioning of close partners. In this regard, providing couple- or household-level intervention frameworks could be effective in achieving better health outcomes for the entire family compared to focusing solely on the depressed individual. Relatedly, existing surveys often collect individual-level data only, and lack information on family members. Understanding familial relationships might elucidate deeper insights into potential effects of depression on other individuals’ health. Furthermore, couples generally share various health-related assets, such as income and dietary habits. Therefore, measurement of couples as a unit in health statistics might provide a more comprehensive measure of population health, which could be further developed into more efficient and comprehensive surveillance and health impact assessment frameworks (Christakis & Fowler, 2009; Miller, 1974).

The present study has three major strengths. First, we performed analyses using a nationally representative survey. Second, we considered a broad array of major characteristics of couples and households, including sociodemographic profiles, physical health, and health behaviors. These adjustments are likely to reduce confounding, particularly bias associated with assortative mating. Lastly, our assessment of depression status was based on validated self-report measures of depressive symptoms completed by index individuals and their spouses independently, which mitigates potential misclassification of their exposure status. We also note two important limitations. First, unmeasured cofounding remains possible, particularly that related to bias associated with assortative mating (Robinson et al., 2017). Second, while exposure status was determined at study baseline, having only a single measurement of depression might cause misclassification. Although the cutpoint used with the CES-D8 score is reported to have reasonably high diagnostic accuracy, potential inclusion of false-positive cases might have biased our results toward null (Steffick, 2000). While depression could also be defined using different cutpoints (e.g., ≥4), our study was constrained in testing alternative cutpoints due to the limited sample size in the exposure subgroups (Dang et al., 2020). Future studies should validate our findings by assessing depression status via sources other than a CES-D8 symptom measure.

In conclusion, the present study examined the separate and joint associations of depression within a couple on an individual’s hazard of mortality. Our findings suggest the harmful effects of one’s depression could extend to the partners’ physical health with consequences for major health endpoints such as mortality. The association between spousal depression and individuals’ mortality was present before the spouse died, indicating widowhood is unlikely to fully explain the observed relationships, and further was maintained after adjusting for a broad range of potential confounders. Future studies on the health effects of depression should incorporate familial contexts, as many have only focused on individual-level characteristics.

## Ethics declarations

All authors state that they have no conflict of interest.

## Author Contributions

T.K. and S.S. had full access to all of the data in the study and take responsibility for the integrity of the data and the accuracy of the data analysis. T.K. and L.K. contributed to the conceptualization and design. T.K. and S.S. extracted and analyzed data. T.K., S.S., and L.K. contributed to the interpretation of results. T.K. contributed to writing the original draft. S.S. and L.K. contributed to reviewing and editing the draft. L.K. supervised the study.

## Data Availability

Data and codebook of HRS are available at https://hrsdata.isr.umich.edu/data-products/public-survey-data upon registration at the website.

