## Appendix for "The Separate and Joint Associations of Own and Spousal Depression with Mortality in Couples"

**Appendix.** Covariate Measurement Details.

**Table S1.** List of Covariates.

**Figure S1.** Sample Flow Chart.

**Figure S2.** Diagram of Multistate Model Analysis.

**Figure S3.** Associations between Depression Status Profile in Couples and Index Individual’s Mortality Adjusted for All Covariates and Including Weights Accounting for Dropped Samples in the Baseline (N = 8,442).

**Figure S4.** Associations between Depression Status Profile in Couples and Index Individual’s Mortality Adjusted for All Covariates for Respondents Completing ≥50% of CES-D8 Items (N = 8,391).

**Figure S5.** Associations between Depression Status Profile in Couples and Index Individual’s Mortality Adjusted for All Covariates for Respondents Excluding Individuals with Proxy Responses for Covariates at Pre-baseline (N = 8,016).

**Appendix.** Covariate Measurement Details

For index individuals, the following covariates were used: demographic (age, gender, race, education, religious denomination, employment, health insurance, and a childhood adversity score), physical health conditions and disability (comorbidity score, overweight/obesity, cognitive impairment/dementia, instrumental activities of daily living (IADL) score), and a composite health behavior score. Childhood adversity was assessed by asking individuals if they experienced each the following experiences in childhood: i) physical abuse, ii) alcohol/drug issue of parents, iii) repeating a year of school, and iv) trouble with police; each experience was scored 1 if individuals reported having it, and then items were summed such that a greater score indicates more adversity (Bürgin et al., 2021). A comorbidity score was derived based on a composite sum score of history of four major chronic diseases: i) heart disease, ii) stroke, iii) diabetes, and iv) cancer (any), where each condition was scored as 0 (no history) or 1 (has history of the disease). The overall score ranged from 0-4, where higher score indicates having a history of more conditions. Cognitive impairment/dementia was assessed using the Telephone Interview for Cognitive Status (TICS, 0-27 points) (Brandt et al., 1988). The TICS score was calculated by summing scores on the four assessments, including immediate recall (0-10 points), delayed recall (0-10 points), two trials of backward counting (0-2 points), and serial 7 subtraction (0-5 points), where higher scores indicate better cognitive function. Following prior work, those with TICS score <12 was classified as having cognitive impairment/dementia (Langa et al., 2017). An IADL score was derived from the composite sum score of 5 items (yes/no; 1 point assigned for reporting difficulty on any given activity): 1) using telephone, 2) managing money, 3) taking medications, 4) shopping for grocery, and 5) preparing hot meal. A composite health behavior score was measured based on the composite sum score of whether repondents met each of three health behaviors (no/yes; 1 point assigned for presence of each unhealthy behavior): 1) binge drinking, 2) smoking, and 3) physical inactivity. Based on a previous study using HRS data (Kim et al., 2021), the characterization of each health behavior was determined as follows: for binge drinking, whether participants had four or more drinks on one occasion in the past 3 months; for smoking, whether they had current smoking habit; and for physical inactivity, whether participants were not engaging in vigorous/moderate physical activity >1 per week. The overall score ranged from 0-3, where higher score indicates having more unhealthy behaviors. We selected the same set of covariates for spouses, except for their childhood adversity score because missing data was more prevalent for this variable, and hence imputation would require stronger assumptions. For household characteristics, we obtained the following: income (in dollars), wealth (in dollars), and length of marriage (in years). All covariates were measured in the pre-baseline (HRS 2008); however, the childhood adversity score for half of the index individuals was derived from the 2010 baseline because HRS collects childhood experiences in every other wave for each half of participants. For all analyses, we standardized age by subtracting the mean age of the analytic sample and log transformed household income and wealth to improve the model fit.

**Table S1.** List of Covariates.

| Variable | Original Measurement | Coding in Analysis |
| --- | --- | --- |
| **Sociodemographic Factors** |  |  |
| Age | Continuous | Continuous |
| Gender | 1= Man; 2 = Woman | 0= Man; 1 = Woman |
| Race | 1= White/Caucasian 2= Black/African American 3= Other  1= Hispanic/Latino  0= non-Hispanic/Latino | 0= White  1= Black  2= Hispanic/Latino  3= Others |
| Educational Attainment | 1= Less than high-school  2= GED  3= High-school graduate  4= Some college  5= College and above | 0= Less than High School  1= High School or Equivalent 2= Higher than High School |
| Religious Denomination | 1= Protestant  2= Catholic  3= Jewish  4= None  5= Others | 0= Protestant  1= Catholic  2= Jewish  3= Others  4= None |
| Household Income ($) | Continuous | Continuous |
| Total Wealth ($) | Continuous | Continuous |
| Employment Status | 1= Works full time  2= Works part time  3= Unemployed  4= Partly retired  5= Retired  6= Disabled  7= Not in Labor force | 0= Unemployed  1= Retired  2= Employed |
| Health Insurance | 0= Not covered by health insurance  1= Covered by health insurance (offered by government plan, employer, or others) | 0= Not covered by health insurance  1= Covered by health insurance |
| Marriage Length (Year) | Continuous | Continuous |
| Childhood Adversity Score | Before age of 18, whether the respondent had…  1) physical abuse by parent.  2) alcohol/drug issues of parent.  3) repeated a school year.  4) trouble with police | Coded as 1 if the respondent reported experiencing an adverse childhood experience, and as 0 if the respondent did not. The total score was derived from the sum of the 4 items, where greater score indicates more adverse childhood experiences. |
| **Physical Health** |  |  |
| Comorbidity Score | Ever had a diagnosis of…  1) heart disease  2) stroke  3) diabetes  4) cancer | Coded as 1 if the respondent reported a diagnosis of each disease, and as 0 if the respondent did not. The total score was derived from the sum of the 4 disease histories, where a greater score indicates a higher number of disease histories. |
| Overweight/Obesity | Self-reported height  Self-reported weight  Body mass index (BMI) was calculated via the formula: weight(kg)/height(m)^2^ | 0= BMI < 25  1= BMI ≥ 25 |
| Cognitive Impairment/Dementia | The Telephone Interview for Cognitive Status (TICS, 0-27 points) was measured based on:  1) immediate word recall (0-10 point)  2) delayed word recall (0-10 point)  3) backward counting (0-2 point)  4) serial 7 (0-5 point) | 0= TICS score ≥12  1= TICS score <12 |
| Instrumental Activities of Daily Living (IADL) Score | Because of a health or memory problem do you have any difficulty with…  1) making telephone call  2) managing money  3) taking medications  4) shopping for grocery  5) preparing hot meal | Coded as 1 if the respondent reported experiencing a difficulty, and as 0 if the respondent did not. The total score was derived from the sum of the 5 items, where greater score indicates more difficulties. |
| **Health Behaviors** |  |  |
| Composite Health Behavior Score | 1) currently smoke (yes/no)  2) whether ever had alcohol drinks; and if yes, the number of days participants had four or more drinks on one occasion in the past 3 months  3) for vigorous/moderate physical activity:  1= Every day  2= >1 per week  3= 1 per week  4= l -3 per month  5= Never | Coded as 1 if the respondent had each unhealthy behavior, and as 0 if the respondent did not. The total score was derived from the sum of the 3 items, where greater score indicates having more unhealthy behaviors.  1) smoking:  0= Does not currently smoke  1= Currently smokes  2) binge drinking:  0= Did not have four or more drinks on one occasion in the past 3 months  1= Had four or more drinks on one occasion in the past 3 months  3) physical inactivity:  0= Engaging in vigorous/moderate physical activity >1 per week  1= Not engaging in vigorous/moderate physical activity >1 per week |

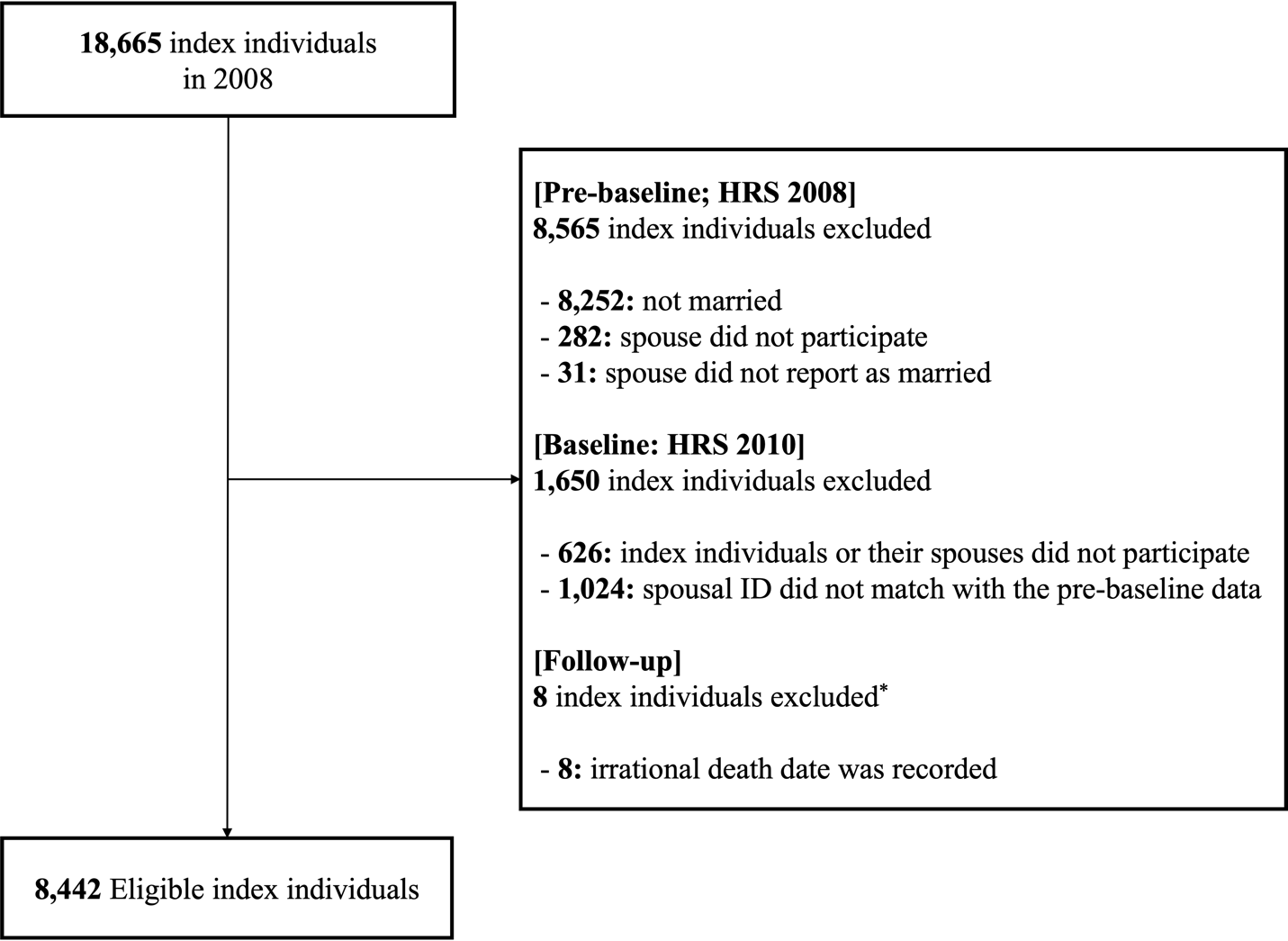

**Figure S1.** Sample Flow Chart.

*The index individuals, whose death dates were recorded in irrational time (i.e., before the baseline interview), were excluded from the analytic sample. However, their data were still leveraged in the analysis as spousal information when their spouses were assigned as index individuals.

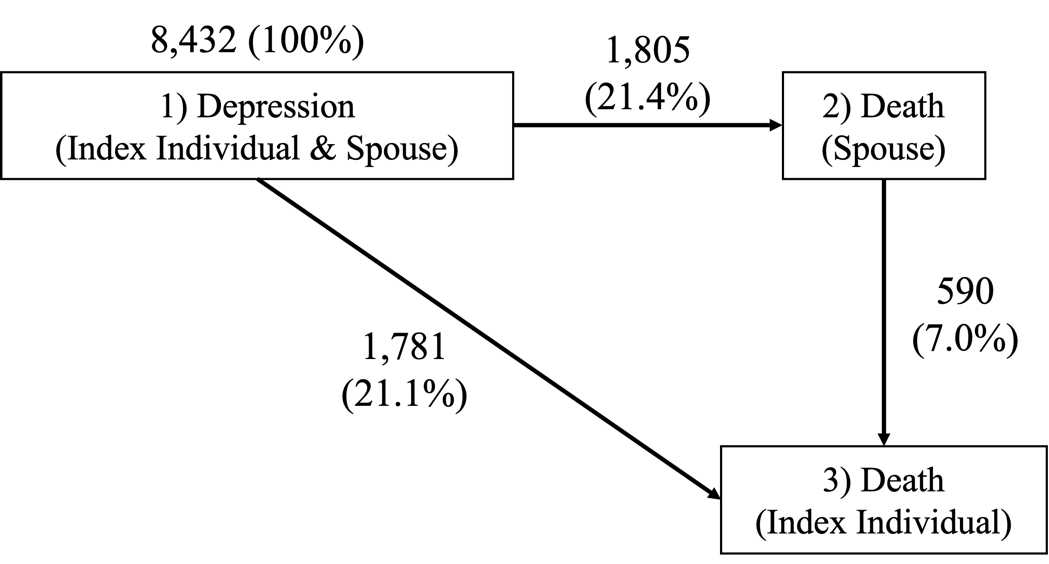

**Figure S2.** Diagram of Multistate Model Analysis.^*,†^

*The periods between 1) depression (index individual & spouse) and 3) death (index individual) was used to estimate the mortality risks associated with exposure status before a spouse’s death.

†The periods between 2) death (spouse) and 3) death (index individual) was used to estimate the mortality risks associated with exposure status after a spouse’s death.

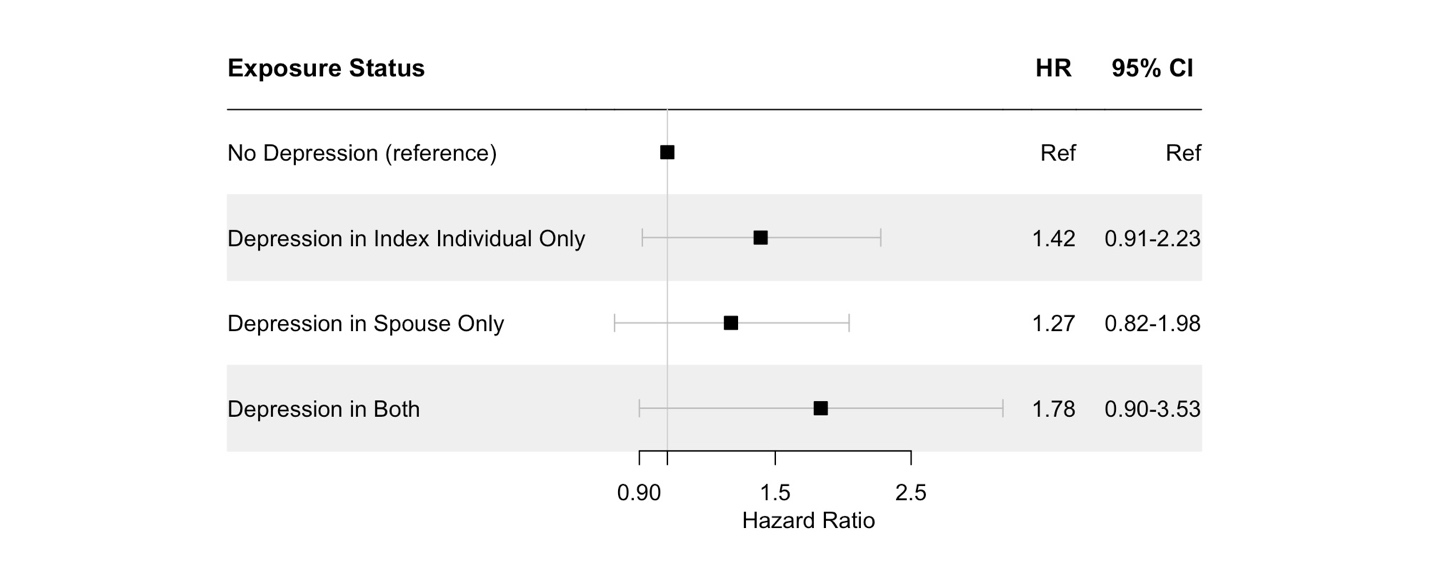

**Figure S3.** Associations between Depression Status Profile in Couples and Index Individual’s Mortality Adjusted for All Covariates and Including Weights Accounting for Dropped Samples in the Baseline (N = 8,442).^*^

Abbreviations: HR, hazard ratio; and CI, confidence interval.

*Hazard ratios were estimated via Cox proportional hazards model adjusted for characteristics of index individuals (age, gender, race, education, religious denomination, employment, health insurance, childhood adversity score, comorbidity score, overweight/obesity, cognitive impairment/dementia, IADL score, and health behavior score), spouses (age, gender, race, education, religious denomination, employment, health insurance, comorbidity score, overweight/obesity, cognitive impairment/dementia, IADL score, and health behavior score), and household (income, wealth, and the length of current marriage).

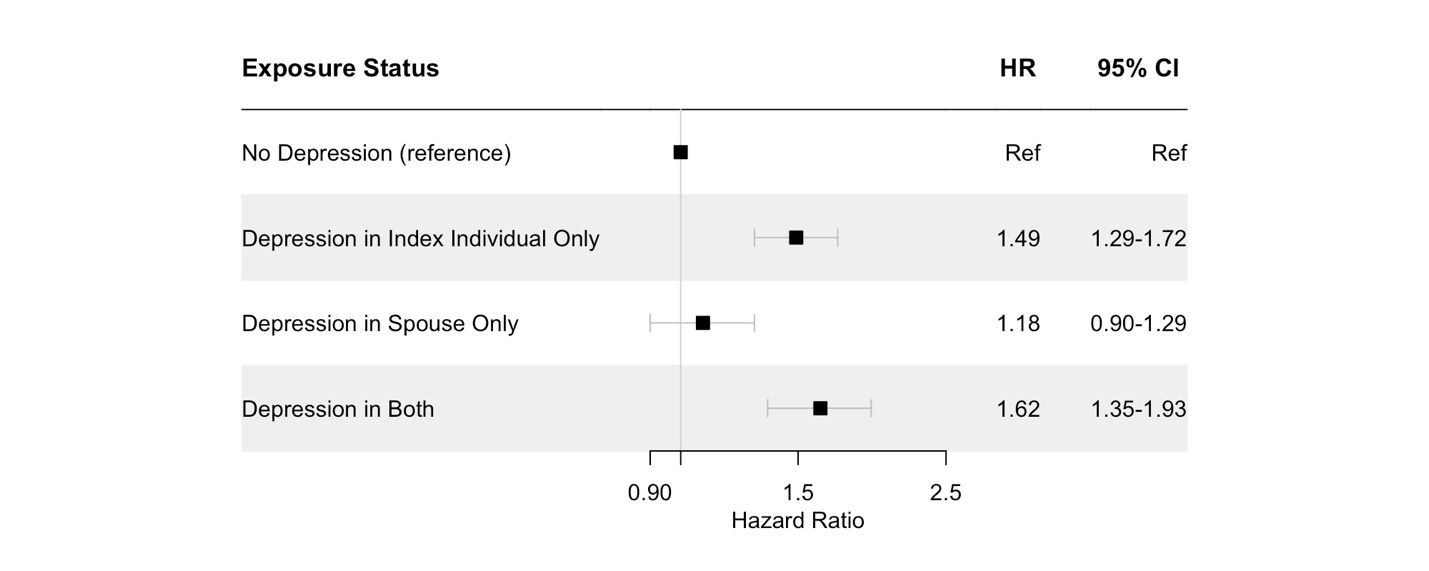

**Figure S4.** Associations between Depression Status Profile in Couples and Index Individual’s Mortality Adjusted for All Covariates for Respondents Completing ≥50% of CES-D8 Items (N = 7,296).^*^

Abbreviations: HR, hazard ratio; and CI, confidence interval.

*Hazard ratios were estimated via Cox proportional hazards model adjusted for characteristics of index individuals (age, gender, race, education, religious denomination, employment, health insurance, childhood adversity score, comorbidity score, overweight/obesity, cognitive impairment/dementia, IADL score, and health behavior score), spouses (age, gender, race, education, religious denomination, employment, health insurance, comorbidity score, overweight/obesity, cognitive impairment/dementia, IADL score, and health behavior score), and household (income, wealth, and the length of current marriage).

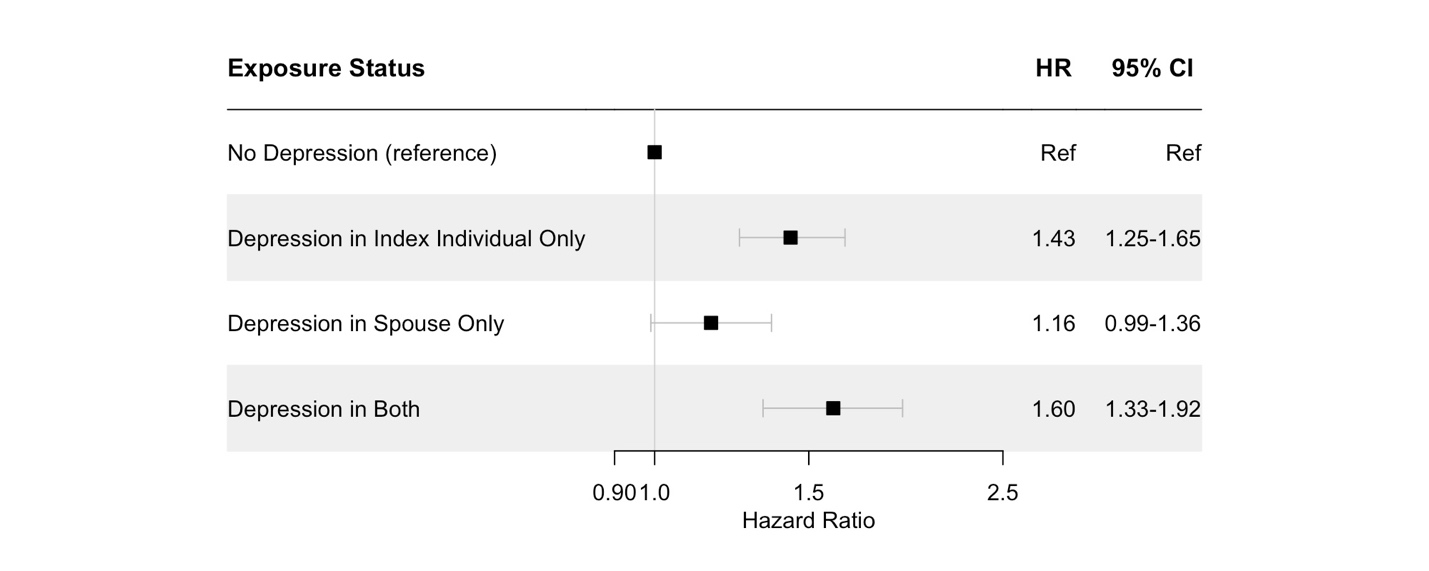

**Figure S5.** Associations between Depression Status Profile in Couples and Index Individual’s Mortality Adjusted for All Covariates for Respondents Excluding Individuals with Proxy Responses for Covariates at Pre-baseline (N = 8,016).^*^

Abbreviations: HR, hazard ratio; and CI, confidence interval.

*Hazard ratios were estimated via Cox proportional hazards model adjusted for characteristics of index individuals (age, gender, race, education, religious denomination, employment, health insurance, childhood adversity score, comorbidity score, overweight/obesity, cognitive impairment/dementia, IADL score, and health behavior score), spouses (age, gender, race, education, religious denomination, employment, health insurance, comorbidity score, overweight/obesity, cognitive impairment/dementia, IADL score, and health behavior score), and household (income, wealth, and the length of current marriage).
